# Time use and social mixing during and around festive periods: Potential changes in the age distribution of COVID-19 cases from increased intergenerational interactions

**DOI:** 10.1101/2020.12.21.20248607

**Authors:** Edwin van Leeuwen, Frank G. Sandmann, Rosalind M. Eggo, PHE Joint modelling group, Peter J. White

## Abstract

**Rationale:** Amid the ongoing coronavirus disease 2019 (COVID-19) pandemic in which many countries have adopted physical distancing measures, tiered restrictions, and episodic “lockdowns,” the impact of potentially increased social mixing during festive holidays on the age distribution of new COVID-19 cases remains unclear.

**Objective:** We aimed to gain insights into possible changes in the age distribution of COVID-19 cases in the UK after temporarily increased intergenerational interactions in late December 2020.

**Method:** We modelled changes in time use and social mixing based on age-stratified contact rates using historical nationally-representative surveys and up-to-date Google mobility data from four weeks before and after the festive period. We explored changes in the effective reproduction number and the age distribution of cases, in four scenarios: (1) “normal”: time use and contact patterns as observed historically, (2) “pre-lockdown”: patterns as seen before the lockdown in November 2020, (3) “lockdown”: patterns restricted as in November 2020, and (4) “festive break”: similar to 3 but with social visits over the holiday period as in 1.

**Results:** Across ages, the estimated *R*_eff_ decreases during the festive break in scenarios 1-3 and returns to pre-holiday levels in scenarios 2-3, while remaining relatively stable in scenario 4. Relative incidence is likely to decrease in children aged 0-15 but increase in other ages. Changes in age distribution were large during the holidays, and are likely to start before the holidays for individuals aged 16-24 years in scenarios 1-3.

**Conclusions:** Our modelling findings suggest that increased contacts during the festive period may shift the age distribution of COVID-19 cases from children towards adults. Given that COVID-19-related hospitalisations and deaths rise by age, more intergenerational mixing risks an increased burden in the period following the holidays.

**Highlights:**

- Home visits are associated with increased intergenerational mixing.
- The effective reproduction number is likely to remain stable or even reduce slightly due to a reduction in contacts at work and school.
- Relative incidence is likely to become lower in children, but higher in the
- older (more vulnerable) age groups around the holiday period, which could lead to increased health care burden.

## Introduction

In response to the newly emerged coronavirus disease 2019 (COVID-19), non-pharmaceutical interventions (NPIs) like physical distancing have been adopted repeatedly in many countries throughout 2020 (Li et al. 2020). These public health measures aim to reduce the close-contact mixing of individuals to reduce or stop the transmission of infectious diseases (Read et al. 2012). Introducing NPIs has been estimated to result in an overall reduction of new COVID-19 cases of 13% (95% confidence interval: 11%-15%) in 149 countries (Islam et al. 2020).

In the UK, a strict national “lockdown” was first implemented in late March 2020, including closure of schools, with some variation in the speed of easing restrictions across the four UK nations between May-July 2020 (Cameron-Blake et al. 2020). Another national lockdown was implemented in England from 05 November to 02 December 2020 that saw schools and universities remain open (Foundation 2020). This was followed by different levels of regionally-targeted restrictions. During the initial national lockdown, residents in England reduced the number of contacts and time spent on activities with higher risk of infection by 48%-74% as compared to what was observed historically (Del Fava et al. 2020; Jonathan Gershuny et al. 2020; Jarvis et al. 2020). Consequently, the lockdown was successful in reducing the estimated effective reprodcution number (*R*_eff_) in England by 75% from 2.6 to 0.61 (Birrell et al. 2020). The effects of the second national lockdown and subsequent regional restrictions are not yet known; however, temporarily lifting some of the restrictions in late December 2020 has been discussed given the “desire to socialise over Christmas” (Williams et al. 2020). For instance, the UK planned to allow up to three households to stay together and form an exclusive “Christmas bubble” ((Public Health England 2020)). This guidance has now been changed in parts of the UK, where a stronger lockdown is being enforced.

Throughout 2020, countries have restricted major festive and religious celebrations that fell in the periods of enhanced physical distancing and lockdowns, including for the Lunar New Year in China at the beginning of the pandemic in January 2020 (Prem et al. 2020). The rationale for (keeping) restrictions during festive and religious periods is the change in social mixing patterns of populations, with more time spent on close-contact interactions between individuals of different ages and from different households. Major religious events have also been associated with an exponentially increasing number of COVID-19 cases both at the initial stages of the pandemic (as seen e.g. in Israel in early March 2020 where *R*_eff_ increased from an estimated 0.69 before a religious holiday to 2.19 afterwards; (Klausner et al. 2020)) as well as later in the year (e.g. two major religious events in Bahrain in September 2020 led to an observed increase in COVID-19 cases of 67% and 119%, respectively; (Abdulrahman et al. 2020)). More generally, lifting NPIs in 131 countries in 2020 was associated with an increase in *R*_eff_ of 11%-25% after 28 days (Li et al. 2020); it took a median of 17 days (interquartile range: 14-20 days) to see 60% of the maximum increase in *R*_eff_ (Li et al. 2020).

Increased intergenerational contacts during festive and religious periods – either encouraged by lifting restrictions; or by individuals breaking rules or interpreting them most favourably (Williams et al. 2020) – risk an relative increase in cases in the older adults, who are at greater risk of severe illness. This could result in an increased burden of COVID-19 after the holidays. Therefore, we aimed to gain insights into possible changes in the age distribution of new COVID-19 cases in the UK after hypothetical temporarily increased intergenerational interactions in December 2020.

## Materials and Methods

For this study we explored how time spent on different activities and in different settings changed over the year. We grouped all activities in the UK time use survey by individual and according to the main activity codes (J. Gershuny and O. Sullivan 2017). These codes were then associated with certain locations that matched the ones in POLYMOD (a large-scale social contact survey; locations included “home,” “leisure,” “school,” “work,” “otherplace,” and “transport”) as well as with certain activities (e.g. social visits, bar/cafe/restaurant visits, park visits, non-essential and essential shopping; for a full list of codes see Table S1 in the supplementary material of van Leeuwen, PHE Joint modelling group, and Sandmann (2020)). We bootstrapped the individual respondents per week, age group, and week day in 5,000 iterations to obtain more robust estimates of the mean time spent on activities within 24 hours (yet we caution about some of the small numbers when stratifying by age group and week, which is reflected in the rapid weekly changes in some of the activities).

For this particular analysis we further grouped the data by two week intervals around the Christmas period from four weeks before to four weeks after the festive period (i.e. end November to early February) and looked at the changes in time use between those two week intervals. Furthermore, we grouped the activities by associated POLYMOD location (van Leeuwen, PHE Joint modelling group, and Sandmann 2020) and normalised the time use by location around the mean. The number of contacts by each location are then scaled according to this normalised value to calculate changes in contacts by location over time. To account for uncertainty in the contact survey data, the POLYMOD dataset was bootstrapped using the socialmixr R package. The “home” location was split into contacts with household members and visitors based on age of the participant, contact and household members (see Supplementary Information). Finally, we defined a number of different scenarios (Table 1) and performed a spectral analysis of the resulting matrices to explore likely changes in transmission and age distribution over the holiday period.

**Table 1:** Different scenarios explored using the spectral analysis. Activity levels are matched to different Google mobility categories. If no clear match exists then the Retail and Recreation category is used as a proxy. Different scenarios use Google mobility from different dates. The festive break scenario is as the lockdown scenario, but assuming social visits return to historic levels during the Holidays

| Scenario | Reference date | Description |
| --- | --- | --- |
| Normal | NA | Using all contacts in POLYMOD |
| Prelockdown | 2020-10-12 | Scaling activities by google mobility levels before the lockdown |
| Current | 2020-11-16 | Scaling by recent levels |
| Festive break | 2020-11-16 | Scaling by recent levels, except for visiting rates over the Hol., which are based on historic rates |

### Datasources

Time-use surveys are used internationally to collect information on how much time individuals spend per day on a wide range of social activities, at what location, and with some limited information on with whom (Bauman, Bittman, and Gershuny 2019). We used individual time-use data from the UK from 2014-2015 with the United Kingdom Time Use Survey (UKTUS) (J. Gershuny and O. Sullivan 2017), which was a nationally-representative survey that collected data on the frequency, duration, and location of a wide range of daily activities in 16,550 diary entries of 9,388 individuals aged 8+ years, and whether activities were spent alone or with others. Participants were asked to complete diaries for 24 hours on two randomly selected days (one on a weekday, one on a weekend day), recording sequences of activities at intervals of 10 minutes (J. Gershuny and O. Sullivan 2017).

The number of contacts by location were based on the POLYMOD survey (Mossong et al. 2008). This survey included data from 1012 participants from the UK with 11882 unique contacts. Participants from all age groups were included. We used publicly available Google mobility data (“COVID-19 Community Mobility Report,” n.d.) that we matched to activity levels, such as work, transport and park visits. For most other activities no clear match existed and in that case the “Retail and Recreation” category was used as a proxy. Time wise (and contact wise) the most important activity in this category was social visits. We used data from different weeks to inform two different mobility scenarios that reflected either the period before the second lockdown in England in November-December 2020 (week starting 12 October 2020) or the period of within the second lockdown (week starting 16 November 2020).

### Incorporating time use into contact matrices

Following previous work (van Leeuwen, PHE Joint modelling group, and Sandmann 2020), we used the following derivation to calculate contact weight:

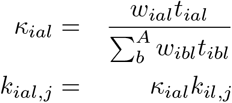

with *κ*_*ial*_ the activity weight for age group *i*, activity *a* at location *l, t*_*ial*_ is the average time spent by an individual, *A* is the set of activities and *w*_*ial*_ is an activity specific weight, which reflects the relative number of people met during this activity compared to other activities at the same location. In the absence of more detailed data (and following our previous work) weights will be set to 1, except for activities that have no new contacts associated with them (i.e. sleep and alone time), which were given a weight of 0 (van Leeuwen, PHE Joint modelling group, and Sandmann 2020).

To account for changes over the year in time spent we added an additional scaling, to scale location contacts with the relative amount spent in those locations. The relative amount of time spent in each location, is included as below.

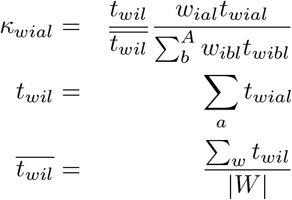

where *t*_*wil*_ is the time spent that period (*w* ∈ *W*) by age group *i* in location *l* and 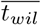 is the time spent by age group *i* in location *l* averaged over all periods.

### Spectral analysis

To understand how the time use and contact pattern changes over the period influence the epidemical model we performed a spectral analysis. The spectral analyis decomposes the dynamical system into its eigenvalues and eigenvectors. In an age stratified SIR model 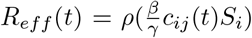, where *ρ* indicates the dominant eigenvalue, *β* and *γ* are, respectively, the transmissibility and the rate of recovery. A typical infected will then be defined by the eigenvector (*v*) associated with the dominant eigenvalue at a given time (*t*). Here the typical infected is the probability that an infected is part of a certain age group/compartment (Diekmann et al. 1990; *τ* (*i*) = *v*_*i*_*/ ∑*_*j*_*v*_*j*_). This value is independent of the exact value of *β* and *γ*. Note that for this analysis we assumed household contacts were stable, to account for the fact that more time at home would lead to higher number of interactions, but not to more unique contacts.

## Results

Timeuse as measured in the timeuse survey changed around the Christmas period (Figure 1). In general, most time is spent at home, school or work and social visits. A substantial amount of time is also spent sleeping, but following previous work we assume that sleep does not lead to any new unique contacts (van Leeuwen, PHE Joint modelling group, and Sandmann 2020). Other highlighted categories are the (non-essential) shopping and bars and restaurants, because these are of particular interest around Christmas. Most age groups spent more time at home over Christmas, except for the elderly. The 16-24 age group spent the least amount of time at home in the period leading up to Christmas. This age group also shows a marked increase in time spent on social visits the 2 weeks leading up to Christmas on during the Christmas holiday. Other age groups spent more time on visits during Christmas, but the change is less pronounced. Patterns for (non-essential) shopping and bars and restaurants differed by age group. Time spent working and at school are reduced over Christmas, although less pronounced in the older age groups, than in the younger groups.

**Figure 1:**
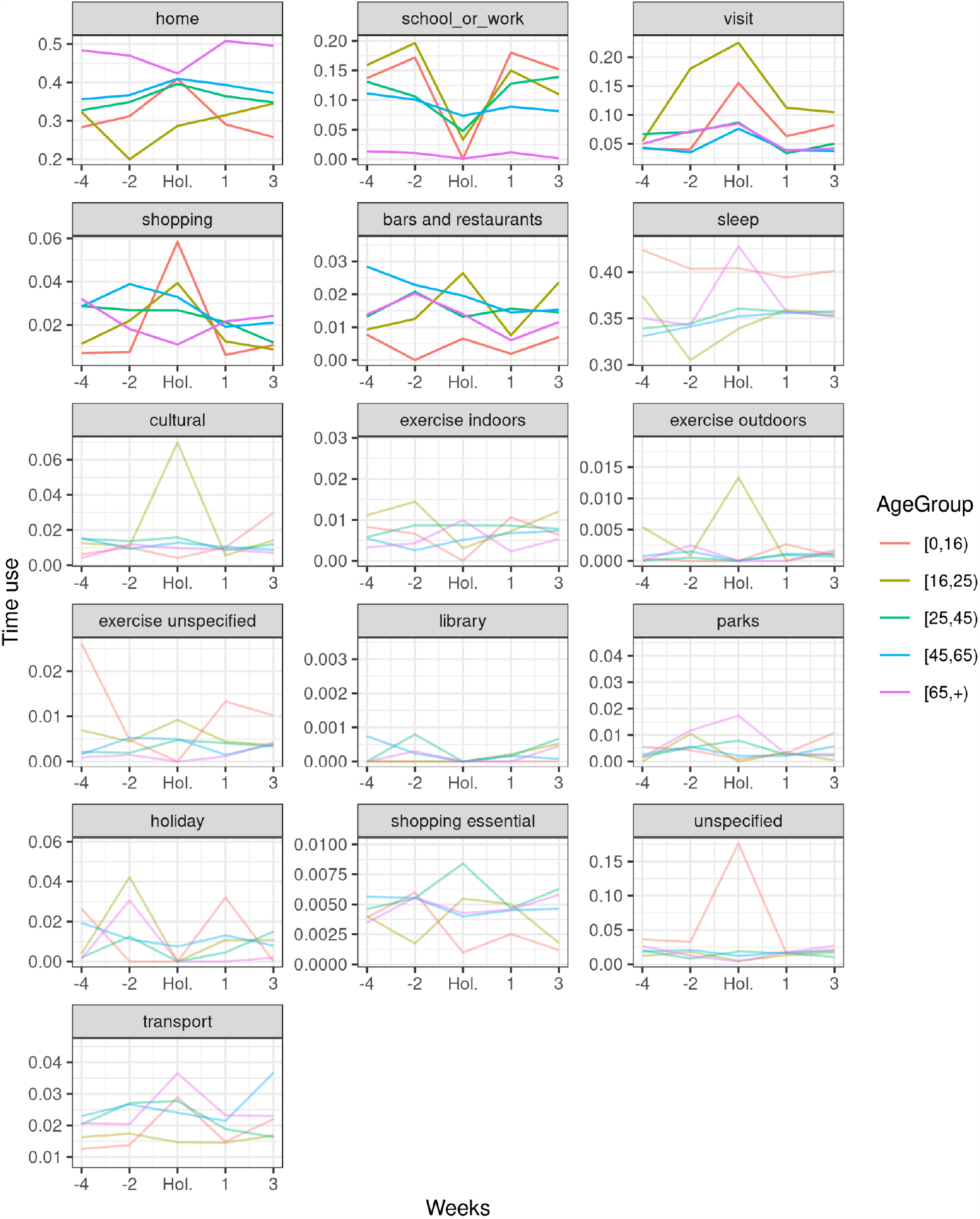
Time use changes around Christmas by activity. Top activities by time spent (except for sleep, which does not lead to new contacts) and activities that are of special interest over the Christmas holiday (e.g. (christmas) shopping and bars and restaurants) are shown in the top rows (other activities are muted in colour, to ease readability). Most people spend more time at home during the Christmas holidays, except for the elderly and the [16, 25) age group. People spend less time at school or work, although this effect is less strong in adults than in children and young adults. Much more time is spent visiting people, especially in the [16, 25) age group. For (retail) shopping and bars and restuarants the patterns are less clear, except that both are clearly higher for the [16, 25) age groups.

Social visits are an important source of contacts, which, based on time use, changes over the Christmas holidays. To understand how these changes could influence transmission it is crucial to understand the age distribution of contacts during such visits. For visits outside of the home we assumed that the POLYMOD contacts distribution during social visits is similar to contacts identified at leisure time in general, but for inside the home we cannot easily do this. Instead we split the contacts into household specific contacts and external contacts. These external contacts were assumed to represent the contact distribution during social visits. Based on this method it is clear that the age distribution of external contacts is different than the distribution of contacts with household members only (Figure S1). Particularly during home visits we would expect many more contacts between elderly individuals and the rest of the population. This is in line with the fact that most over 65 live with other over 65, but not with younger generations.

We found clear increases in contacts made during visits at home in all age groups, which are associated with high intergenerational mixing (Figure S1 and S2). This increase was particularly high in the 16-24 age group, were it started two weeks before the holiday (Figure S2). In contrast, school and work related contacts were reduced across all age groups. The reduction in “school” contacts for the youngest age group is very high, but this is partly offset by increased contacts at other locations. The 16+ age group also had reduced contacts due to closing of schools and reduced time spent working, but especially for the 16-24 age group this is offset by the increase contacts related to visits.

Spectral analysis showed that the changes to the underlying dynamic system depend on the scenario assumptions, but some general patterns emerge. The dominant eigenvalue is lower during the holidays for the prelockdown and lockdown scenario (Figure 2). This is because in those scenarios contacts outside of school (and to a lesser extent work) were reduced and, therefore, those contacts contribute more to mixing than in the normal scenario. As a result, closing schools and reducing time spent at work had a large effect on the transmission rate. The dominant eigenvalue was actually higher during the holidays in the festive break scenario, because the increase in contacts during visits offset the lost contacts due to closing schools and reduced work contacts.

**Figure 2:**
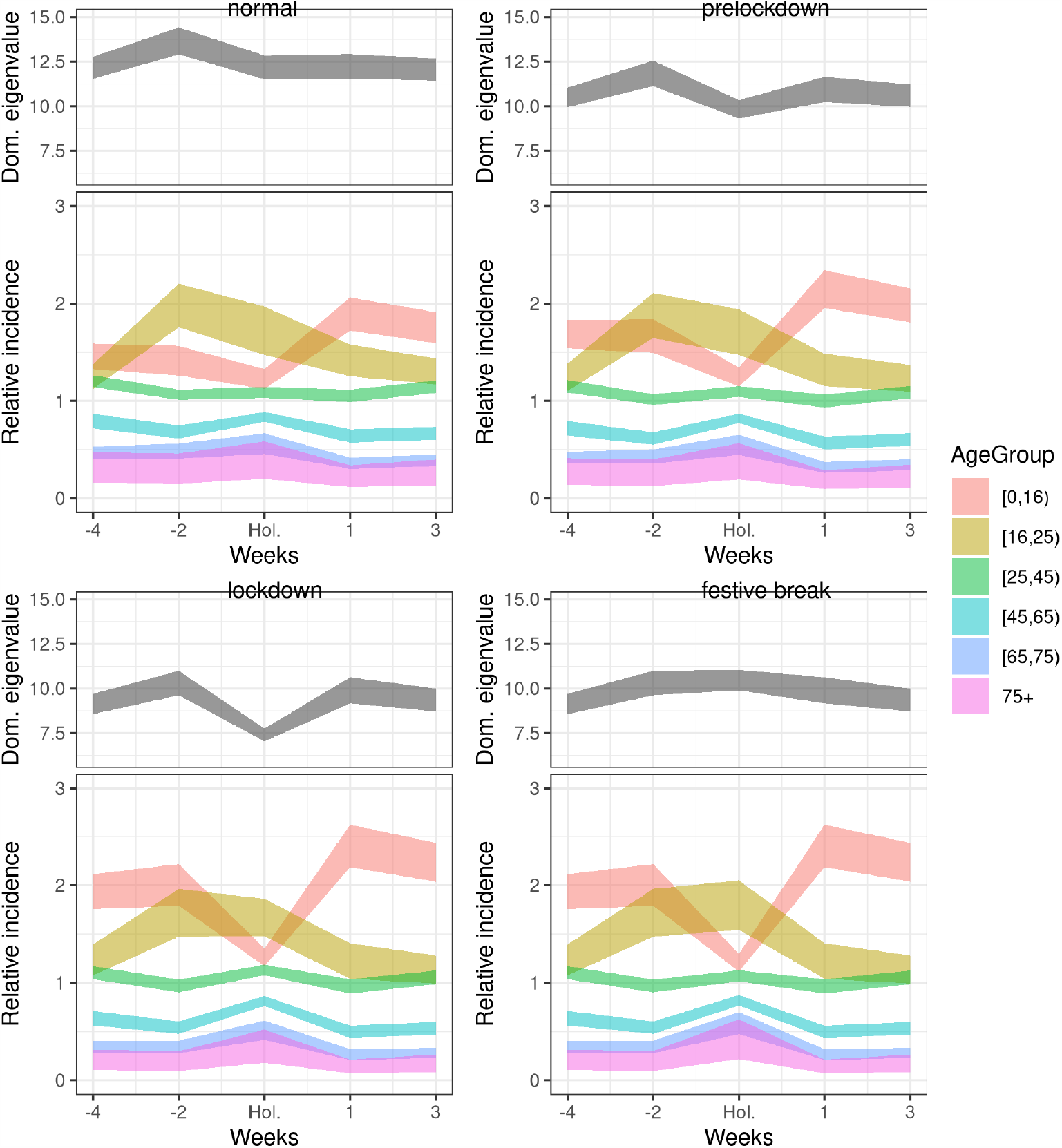
Spectral analysis of the resulting contact matrices for different scenarios. For each scenario the top panel shows the dominant eigenvalue. Relative changes in the eigenvalue directly linked to relative changes in reproduction number (*R*). The bottom panel shows the population weighted eigenvector, which is closely linked to the relative incidence in each age group. The dominant eigenvalue shows different patterns depending on the scenario. The relative incidence in children would likely go down due to reduced contacts, especially those made at school, while for the [16, 25) age group is likely to increase. All other age groups also show a relative increase over the holiday period, indicating that the age distribution of cases is likely to switch from children to the other age groups. For this analysis household contacts were kept constant, based on the assumption that they will be with the same people from day to day.

The eigenvector analysis also highlighted subtle differences between the scenarios, but qualitatively patterns were comparable (Figure 2). Over the holidays we would expect the relative number of infected to go down in the youngest age group, due to closing schools. For the 16-24 age group, relative number of infected would actually be expected to start to go up before Christmas, and remain high over Christmas, mainly driven by an increase in social visits over this time. Finally, the other age groups showed an increase over the holiday, which is particularly marked in the elderly, where it could be doubled. Note though that the eigenvector analysis gives us the attractors of the dynamical system, which indicates how the model converges, but the exact behaviour would be highly dependent on other parameters, such as generation time of the disease.

## Discussion

This modelling study provides insights into the expected changes of transmission and the age distribution of COVID-19 cases before during and after increased intergenerational social mixing. Despite the success of NPIs in reducing the widespread community transmission during the COVID-19 pandemic (Islam et al. 2020; Li et al. 2020), discussions about lifting restrictions have been ongoing in many countries throughout 2020. Our study showed that while the reproduction number may remain relatively stable or even show a slight reduction when relaxing restrictions for a short period of time in December 2020, the age distribution of COVID-19 cases is likely to change from children towards adults. Increased intergenerational contacts during festive periods thus risk a rising burden in terms of morbidity and mortality in the weeks after the holidays given that COVID-19-related hospitalisations and deaths rise by age (Birrell et al. 2020; Docherty et al. 2020).

Although our study resulted in modest changes in reproduction number (and associated growth rate), they appear to be in line with what was observed for other major religious events in 2020 (Abdulrahman et al. 2020; Klausner et al. 2020). Our modelling results also showed that for individuals aged under 16 years a return to school after the holidays could result in a rise in the relative incidence to pre-holiday levels. Furthermore, in some of the scenarios the changes in incidence for individuals aged 16-24 years were largest in the weeks just before the holidays, due to increased visits. The historic increase of social visits in this age group may be reflective of university students usually leaving dormitories and returning to their parents’ homes in the weeks before the festive period. The more staggered approach this year of students leaving dormitories already in early-/mid-December, and returning on campus over a longer period of time in early 2021, may have resulted in prolonged, but much reduced increased mixing in this age group.

### Strengths and Limitations

Our study used nationally-representative survey data and population-level mobility data in a transmission-dynamic compartmental model to explore changes in behaviour and the impact on new COVID-19 cases. This work built on our previous work (van Leeuwen, PHE Joint modelling group, and Sandmann 2020), and preliminary findings were presented to UK scientific advisers to highlight possible scenarios for the festive period in December 2020. In the absence of being able to robustly predict the behaviour of individuals during a major festive holiday in the UK, our study needed to make assumptions about time use and its effect on contact patterns that reflect the historic survey data. The mapping from time use to number of contacts assumes that time use is proportional to number of contacts during that activity, which may not hold for specific settings such as during social visits and in bars and restaurants. Also, our analysis did not explicitly capture the effect of individuals moving between geographical locations and/or potentially changing the composition of their social circles over Christmas, or the potential effects of forming social bubbles over the holiday period. Similar changes have been reported during the summer holidays in the UK in 2020, with Facebook population movement decreasing in more densely populated areas and increasing in rural areas that are popular holiday destinations; however, the movement in areas of localised lockdowns decreased slightly (Gibbs et al. 2020).

No direct data on the age composition of visits over Christmas is available. Instead this study used the “leisure” and “homevisit” data extracted from the POLYMOD survey (Mossong et al. 2008), which represent a broader time period than the Christmas period. Traditionally, Christmas is a holiday focussed on family visits than visits with friends (which are more common outside Christmas period; (Johnes 2016)). As a result intergenerational mixing over this period is likely to be even more pronounced than estimated in this study. This study might therefore underestimate the additional burden in the older generation. However concern about COVID-19 is likely to cause many people to have fewer contacts with their over 65 relatives than in a normal festive period, which could offset this underestimation.

Although we are in the midst of an ongoing pandemic with physical distancing measures having been implemented for an extensive duration, the adherence of individuals with NPIs over the festive period is unclear given the “desire to socialise over Christmas” (Williams et al. 2020). A recent survey reported that support for Christmas guidance was 51% (ONS, n.d.). This may be regarded as relatively low, but even if only half the populaton restricts contacts then this would represent many fewer contacts than in a normal festive period. With the new, stricter guidance coming in for large parts of England and Wales this would lower the number of contacts even further.

It is important that surveys like CoMix are conducted over Christmas so that we can anticipate the likely increase in cases based on knowledge of the actual contact patterns. After the holidays and to curb any projected rise in burden, more restrictive NPIs at population-level may become temporarily necessary again to reduce the community transmission in the UK (Keeling et al. 2020). With the rollout of mass COVID-19 vaccination having just started in the UK in early December 2020 as the first country globally (Mahase 2020a), and full protection of the vaccine taking up to 28 days (Mahase 2020b), physical distancing will remain pivotal in the upcoming festive holidays both in the UK as well as internationally.

## Conclusions

Our modelling findings suggest that increased contacts during the festive period may shift the age distribution of COVID-19 cases from children towards adults. Given that COVID-19-related hospitalisations and deaths rise by age, more intergenerational mixing during and around the festive period risks an increased burden in the time following the holidays.

## Data Availability

All data used in the manuscript has been published before.

## Acknowledgement

We gratefully acknowledge the access to the data from the United Kingdom Time Use Survey through the UK Data Service (http://doi.org/10.5255/UKDASN-8128-1).

