## Supplementary Information for "Time use and social mixing during and around festive periods: Potential changes in the age distribution of COVID-19 cases from increased intergenerational interactions"

### Supplementary material

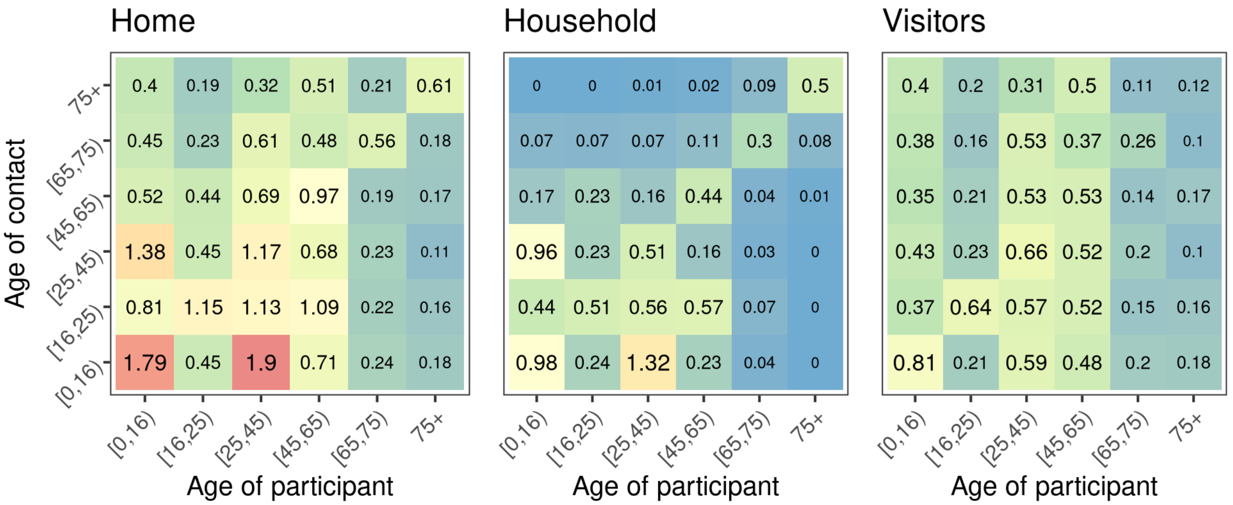


Figure S1: Home contacts were split into household contacts and contacts with visitors. Visitor contacts were defined as contacts where the contact's age is different than any of the ages of the household members. Home household contacts show very little mixing between the under 65 and the over 65. Most mixing is either within the under 65 or within the over 65. The visitor matrix shows contacts across the whole age range.

Home contacts were stratified into visitor and household contacts, by checking the age of the participant, the contact and the household members. If the age of the participants is not equal to the contact's age, but is equal to one of the household members then it was counted as a household contact. Note that this is likely to result in some visitor contacts being associated with household contacts, because they have the same age as one of the household members. If the contact's age is the same as the participant's age, then we check if there are multiple members in this household with that age, if so then we give it a probability that it is a household contact weighted by the number of household members with this age ($1 - 1/n$; where $n$ is number of household members with that age). This was only the case for a fraction of home contacts ($\approx 0.02$) and should have a minimal effect on the final stratification. In all other cases the contact is assumed to be external to the household.

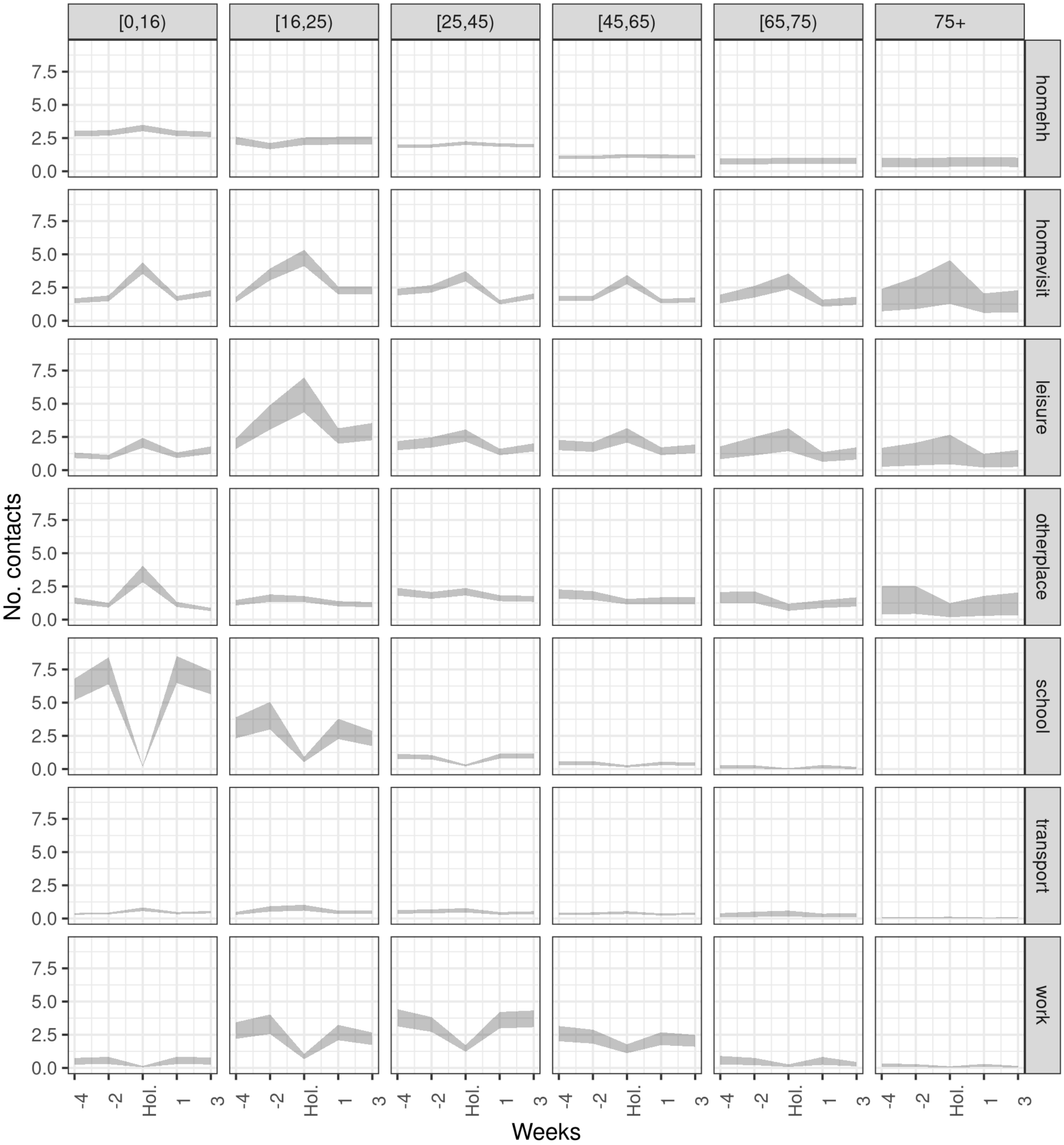


Figure S2: Number of contacts by location and period for different age groups. These figures follow a similar pattern as the previous figure, but important to notice is that the extra contacts for the youngest age group do not offset the number of contacts at school. For the $[16, 25)$ age group visit related contacts during Christmas period are higher than contacts in any other location. For the adult age groups, the main difference is in the reduction in work contacts and increase in contacts associated with social visits.
